# Ultrarare Variants in DNA Damage Repair and Mitochondrial Genes in Pediatric Acute-Onset Neuropsychiatric Syndrome and Acute Behavioral Regression in Neurodevelopmental Disorders

**DOI:** 10.1101/2025.08.27.25333852

**Authors:** Dhanya Vettiatil, Anjana Soorajkumar, Robert A. Dubin, Erika Pedrosa, Allan Schornagel, John S. Lambert, Isadora Pinheiro Costa, Joseph McDonald, Sigrid M.A. Swagemakers, Peter J. van der Spek, Jennifer Frankovich, Janet L. Cunningham, Herbert M. Lachman

**Affiliations:** Department of Psychiatry and Behavioral Sciences, Albert Einstein College of Medicine, Bronx New York, U.S.A; Center for Epigenomics / Computational Genomics Core, Albert Einstein College of Medicine, Bronx New York, U.S.A; Kinderpraktijk Zoetermeer, Zoetermeer, Netherlands; Infectious Diseases Department, Mater Misericordiae University Hospital, UCD School of Medicine, Dublin 7, Ireland; Department of Pediatrics, Section of Pediatric Rheumatology, University of Chicago, Chicago Illinois, U.S.A; Department of Pathology and Clinical Bioinformatics, Erasmus MC, Rotterdam, Netherlands; Department of Pediatrics, Division of Pediatric Allergy, Immunology, Rheumatology and Immune Behavioral Health Program, Stanford Children’s Health and Stanford University School of Medicine, Palo Alto, California, U.S.A; Department of Medical Sciences, Clinical Psychiatry, Uppsala University, Uppsala, Sweden; Departments of Genetics, and Dominick P. Purpura Department of Neuroscience, Albert Einstein College of Medicine, Bronx New York, U.S.A

**Keywords:** PANS, autism, regression, DNA repair, Fanconi anemia complex, neuroinflammatory, neuroinflammation, DUOX2, gut microbiome, Jansen de Vries Syndrome, cGAS-STING, type I interferons, DNA damage response, p53, CCR9

## Abstract

**Introduction:** We recently identified variants in 10 genes that are members of either the p53 pathway or Fanconi Anemia Complex (FAC), regulators of the DNA repair (DNA damage response; DDR) in 17 cases with Pediatric Acute-Onset Neuropsychiatry Syndrome (PANS) or regression in autism spectrum disorder (ASD) and other neurodevelopmental disorders (NDD). We aimed to identify additional cases with genetic vulnerabilities in DDR and related pathways. **Methods.** Whole exome sequencing (WES) and whole genome sequencing (WGS) data from 32 individuals were filtered and analyzed to identify ultrarare pathogenic or likely pathogenic variants.

**Results:** Variants affecting DDR were found in 14 cases diagnosed with PANS or regression (*CUX1, USP45, PARP14, UVSSA, EP300, TREX1, SAMHD1, STK19, MYTl1, TEP1, PIDD1, ADNP, FANCD2*, and *RAD54L*). The *CUX1* variant is de novo, as are two cases who had mutations in genes that affect mitochondrial functions that are connected directly or indirectly to mitophagy (PRKN and *POLG*), which can trigger the same innate immune pathways when disrupted as abnormal DDR. We also found pathogenic or likely pathogenic secondary mutations in several genes that are primarily expressed in the gut that have been implicated in gut microbiome homeostasis (e.g., *LGALS4, DUOX2, CCR9*).

**Conclusion:** These findings align with previous genetic findings and strengthen the hypothesis that abnormal DDR and mitochondrial dysfunction underly pathogenic processes in neuropsychiatric decompensation. The potential involvement of genetic variants in gut microbiome homeostasis is a novel aspect of our study. Functional characterization of the downstream impact of DDR deficits may point to novel treatment strategies.

## Introduction

Abrupt severe psychiatric symptoms and behavioral regression in children are relatively rare and proposed to be linked to underlying immunological deficits [1–3]. One such disorder is Pediatric Acute-Onset Neuropsychiatric Syndrome (PANS), which is characterized by the core symptoms of abrupt onset obsessive-compulsive disorder (OCD) or restricted eating (food aversion), along with two or more secondary symptoms: anxiety, emotional lability, irritability, rage, cognitive decline, sleep disturbance, sensory dysregulation, movement abnormalities, and urinary symptoms including new onset enuresis or urinary frequency [4]. While the criteria require two secondary symptoms, most patients have 5-6 that start abruptly alongside the OCD and/or restricted eating [5–7]. For parents, the abrupt onset is a telling feature, with symptoms usually occurring within a day or two following an infection, with some children showing a rapid decline in hours [4, 8, 9]. During a flare, there is a marked deterioration in school performance, exemplified by an inability to concentrate and the loss of previously learned skills, such as handwriting, artwork, reading, and math, which accompany the severe psychiatric symptoms. Dysautonomia is commonly found, characterized by enuresis, postural orthostatic tachycardia syndrome (POTS), and dilated pupils, which also occurs with joint hypermobility [5, 6, 10, 11].

PANS can occur in children with premorbid autism spectrum disorder (ASD) or other neurodevelopmental disorders (NDDs) [5–7, 10, 12]. The mean age of onset is 6-7 years, although later onset in teenagers and young adults is increasingly being recognized [13–15]. The course is typically relapsing and remitting [5, 16].

As is true with mitochondrial disorders, the part of the brain that is prominently affected in PANS/PANDAS is the basal ganglia (PANDAS is Pediatric Autoimmune Neuropsychiatric Disorders Associated with Streptococcal Infections, a subtype of PANS) [17–20]. Additionally, studies have demonstrated that patients with PANS/PANDAS have autoantibodies that target cholinergic interneurons and dopamine receptors within the basal ganglia [11, 21–24]. These patients also appear to have an increased prevalence of autoantibodies to folate receptors [25]. Autoimmune and other inflammatory disorders are prevalent in first-degree family members [26, 27]. Patients with PANS also have a high rate of enthesitis, arthritis, autoimmune conditions, and other inflammatory/immune disorders [11, 21, 28].

An immune-based etiology is also suggested in ASD and NDD, especially in cases who experience a profound neuropsychiatric deterioration following infection. A strong family history of autoimmune disorders is associated with an increased risk of ASD [29–31]. A striking example is acute and subacute regression triggered by infection and immune stimulation in individuals with certain genetic ASD and NDD subtypes, as *SHANK3*-associated ASD and NDD, and Down Syndrome Regression Disorder (DSRDS) [32–37].

To dissect the underlying molecular and genetic basis of regression and PANS, we have been carrying out whole exome and whole genome sequencing (WES and WGS, respectively). In our original study, ultrarare variants were found in 11 genes in 21 cases [38]. The candidate genes showed extensive heterogeneity but clustered to those that affect innate and adaptive immunity (*PPM1D, NLRC4, RAG1*, *PLCG2,* and *CHK2*), or genes that affect synaptic function (*SGCE*, *CACNA1B*, *SHANK3*, *GRIN2A, GABRG2,* and *SYNGAP1*). Out of the 21 subjects, five were diagnosed with ASD before experiencing a deterioration that met PANS criteria. In addition to ASD, four cases were diagnosed with NDD, one of whom had Jansen de Vries Syndrome (JdVS). JdVS is caused by truncating mutations in *PPM1D* exons 5 or 6 that cause a gain-of-function (GOF) effect by suppressing p53 and other proteins involved in DDR (e.g., MDM2, ATM, CHK1, CHK2, ATR, and H2AX) through an increase in phosphatase activity caused by the loss of a degradation signal located in exon 6 [39–43].

The *CHK2* variant we identified in the study is a frameshift mutation that disrupts the kinase domain and is likely a loss-of-function (LOF) mutation (c1229delC; T410fsX14). Thus, GOF mutations in *PPM1D,* a p53 phosphatase, and LOF mutations in *CHK2,* a p53 kinase, are both expected to disrupt DDR by reducing p53 phosphorylation.

This pathway was supported in our most recent study in which one patient with PANS and a family of siblings with either PANS or ASD deterioration were identified with pathogenic LOF mutations in *ATM,* which like *CHK2*, codes for a kinase that phosphorylates p53 and other DDR proteins [1] [44–48]. Two other JdVS cases who experienced infection-induced neuropsychiatric decline were also presented in the paper, as well as patients with mutations affecting other genes involved in p53-mediated DDR (*ATR, 53BP1*, and *RMRP*). In addition, we identified patients with genetic variants in five members of the Fanconi Anemia Complex (*FANCE, SLX4/FANCP, FANCA, FANCI*, and *FANCC*) [1]. We hypothesized that defects in DNA repair genes, in the context of infection or other stressors, could contribute to decompensated states through an increase in genomic instability with a concomitant accumulation of cytosolic DNA in immune cells triggering DNA sensors, such as cGAS-STING and AIM2 inflammasomes, as well as central deficits on neuroplasticity. Genomic instability could also disrupt immunological responses by adversely affecting mitochondrial function, senescence and apoptosis.

Supporting this hypothesis is a recent report on 41 individuals with DSRD, eight of whom were found to have de novo variants [49]. Among the eight genes, four affect interferon signaling of which three codes for proteins involved in DNA repair.

In addition, in a recent paper by Dale et al., eight children who experienced regression or abrupt-onset neuropsychiatric syndromes following infections were found with pathogenic de novo variants in genes encoding chromatin regulators, the majority of which have DNA repair functions [50].

In this paper, we expand on these genetic findings with continued focus of ultra rare genetic variants that code for proteins involved in DDR. In addition, we report several cases in whom pathogenic mutations were found in genes that affect mitochondrial DNA replication and mitophagy, processes that could also lead to innate immune activation when disrupted. In some cases, secondary mutations were found in genes involved in regulating innate immune function, supporting an oligogenic model for some PANS and regression cases. Notably, we also identified pathogenic variants in genes that are primarily expressed in the gut, including a de novo variant in *CCR9*, a chemokine receptor that plays a key role in gut-associated immune responses. These findings suggest that the gut microbiome, gut-brain axis, and/or gut permeability may be involved in PANS and regression in certain cases.

## Subjects and Methods

### Ethical considerations

This study was conducted in accordance with the Declaration of Helsinki. Informed consent was obtained from all subjects and/or their legal guardian(s). The study was approved by the Albert Einstein College of Medicine Institutional Review Board (IRB; protocol ID 2022-14636).

### Subjects

The subjects were identified through connections between the senior investigators and a PANS group called EXPAND, a non-profit European advocacy organization for families of children, adolescents, and adults with immune-mediated neuropsychiatric disorders (https://expand.care/), through the Neuroimmune Foundation, a non-profit organization dedicated to neuroimmune and inflammatory brain conditions (https://neuroimmune.org/about/), The Louisa Adelynn Johnson Fund for Complex Disease (https://tlajfundforcomplexdisease.com/), the Jansen de Vries Syndrome Foundation (https://jansen-devries.org/), and the Alex Manfull Fund (https://thealexmanfullfund.org/).

Histories were obtained by the participating physicians and confirmed and collated by one of the senior investigators who interviewed the parents of every case (H.M.L). The cases met the criteria for PANS or regression with or without comorbid ASD or NDD. In cases with acute and subacute onset behavioral regression, the loss of previously established skills (such as speech, school performance, executive function, urinary continence, and others) was observed by caregivers and healthcare personnel with a large impact on function. Each case was assigned a study ID that is not known to anyone outside the research group, maintaining patient confidentiality.

### Genetic analysis: Variant calling and annotation

Data from 32 whole exome sequencing (WES) analyses were processed in the Computational Genomics Core at the Albert Einstein College of Medicine after importing sequencing files from GeneDx, Nebula, and CeGaT. Commercially-generated vcf files that had been mapped to human genome hg38 were annotated with Annovar (version: Date: 2020-06-08) [51]; commercially-generated VCF files that had been mapped to hg19 were re-mapped to hg38 using the Picard Tools module LiftoverVcf (version: 2.26.10) prior to annotation. In cases where LiftoverVcf files were not available, fastQ files were aligned to hg38 using the mem algorithm of bwa (version 0.7.17-r1188; https://bio-bwa.sourceforge.net), and bam files were sorted by coordinates, duplicates marked, and base quality scores were recalibrated (Picard Tools modules SortSam and MarkDuplicates; Genome Analysis Toolkit (GATK) v4.4.0.0 modules BaseRecalibrator and ApplyBQSR). Variant calling was performed using Haplotypecaller and raw variants were filtered using modules CNNScoreVariants (1D model, pre-trained architecture) and FilterVariantTranches (GATK, v4.4.0.0); filtered vcf files were annotated by Annovar. In all cases, only entries that passed all filters (PASS in the vcf file FILTER column) were annotated. QC was performed using FastQC, FastQ-screen, and CollectHsMetrics.

For the current study, genes of interest were found in the WES analysis of 10 cases. In two cases, we relied on the analysis carried out by GeneDx (cases 1 and 2). Whole Genome Sequencing (WGS) was carried out on case 11 by one of the co-authors (PS) in the Clinical Bioinformatics Center at Erasmus MC, Rotterdam, Netherlands. In three cases, we relied on findings reported in Invitae panels (Epilepsy, case 12; Autoimmune, case 13), and CeGaT WES trio analysis (case 16).

All ultrarare variants, minor allele frequency (MAF) <0.001 in exons and intron/exon junctions were assessed. MAFs were obtained from the GnomAD database on the UCSC Genome Browser (https://genome.ucsc.edu/). We focused on splice variants, frameshift and non-frameshift mutations, stopgain and stoploss variants, and indels. Nonsynonymous mutations that were predicted to be pathogenic by SIFT and MutationTaster were also evaluated. In addition, all ultrarare variants in ASD and NDD genes found on the SFARI database and Invitae NDD panel were examined. Immune, DNA repair, and mitochondrial genes were evaluated based on those found on Invitae Panels: Immunodeficiency and Autoinflammatory and Autoimmunity Syndromes, DNA Damage Repair, and Nuclear Mitochondrial Disorders.

## Evaluation of nonsynonymous variants

The predicted pathogenicity of each variant based on ACMG criteria (pathogenic, likely pathogenic, uncertain, possibly benign, likely benign) were obtained from Franklin, and in some cases ClinVar and VarSome, with a few exceptions that will be described on a case-by-case basis.

https://franklin.genoox.com/) https://www.ncbi.nlm.nih.gov/clinvar/

https://help.genoox.com/en/articles/6240723-prediction-tools-pp3-bp4 https://varsome.com/

We also used Combined Annotation Dependent Depletion (CADD) scores in assessing single nucleotide variants. CADD provides a Phred-like score that is a measure of deleteriousness [52]. A CADD score represents a ranking, not a prediction, and no threshold is defined. Higher scores are more likely to be deleterious. A score of 20 is commonly used as a cutoff, with scores above 20 predicted to be among the 1.0% most deleterious possible substitutions in the human genome. CADD scores were obtained using a tool developed by the University of Washington, (https://cadd.gs.washington.edu).

Finally, nonsynonymous mutations were also evaluated by AlphaMissense, in which amino acid changes are evaluated by an adaptation of AlphaFold to predict nonsynonymous variant pathogenicity [53]. It combines the structural context of the amino acid substitution and evolutionary conservation to provide a pathogenicity score. AlphaMissense (AM) predicts the pathogenicity of all possible single amino acid substitutions in nearly all protein-coding genes, providing the capacity to classify 89% of nonsynonymous variants.

Splice site variants were evaluated by SpliceAI. A score of 0.5 (recommended) was used as a cutoff (https://spliceailookup.broadinstitute.org/).

### Protein–Protein Interaction Network Analysis

To explore functional relationships among DNA repair gene variants, we performed a protein– protein interaction (PPI) analysis using the STRING database (version 12.0; https://string-db.org/). The analysis included DNA repair genes identified in this and our two previously published studies[1, 38]. The resulting PPI network was exported from STRING and imported into Cytoscape (version 3.10.3; https://cytoscape.org/) for network visualization, layout optimization, and functional annotation. Network modifications and aesthetic enhancements were applied in Cytoscape to generate the figure. Line thickness in the figure is based on a confidence score from 0 to 1, showing how likely STRING thinks a predicted interaction is real. Thicker (heavy) black lines mean higher confidence scores (closer to 1), thinner lines mean lower confidence, which are closer to our cutoff of 0.4 (medium confidence).

### Validation of DNA repair variants

DNA repair variants were validated by Sanger Sequencing (see **Supplemental Methods** for primers) with a few exceptions: the PRKN CNV was validated from WES; Cases 1 and 2 are monozygotic (MZ) twins with the same variant; no DNA available for SAMHD1.

## RESULTS

We identified 14 candidate genes with ultrarare variants that affect DNA repair in 14 cases and an additional 2 cases with mutations in genes that affect mitochondrial DNA (mtDNA) repair or mitophagy, which are included because abnormalities can lead to activation of the innate immune system (described below). The ultrarare variants are predicted to be pathogenic or likely pathogenic, based on VarSome, AM, Franklin criteria, with a few exceptions that will be described below (**Table and Supplemental Table 1**). Most of the DNA repair genes have an effect on innate immunity. We also included two cases with mutations in genes that affect mitochondrial DNA (mtDNA) repair or mitophagy, since abnormalities can lead to activation of the innate immune system (described below). Among the variants, six are nonsynonymous (including two different *CUX1* variants), three disrupt canonical splice sites, four are stopgain variants, there are two frameshifts, and one copy number variant (CNV). Three of the primary variants are de novo (*CUX1* in the MZ twin pair, the *PRKN CNV,* and *POLG*). We could not determine the transmission pattern in one case due to unavailable parental DNA. The remainder were transmitted. There was a strong family history of autoimmune/autoinflammatory disorders in all but one case (case 15). Although a history of autoimmune and autoinflammatory disorders has been observed in PANS families, the extraordinarily high rate in our cohort suggests a selection bias for genes involved in DNA repair and mitochondrial homeostasis. Note that family history details in this report are omitted to maintain confidentiality; researchers can contact the corresponding author for access to those data.

The following is a short clinical description of the cases along with the DNA repair genetic variants we identified, which are listed in the **Table (see Supplemental Table 1** for a more detailed assessment of each variant shown on the **Table**).

### Cases 1, 2, and 3: *CUX1*

The most compelling genetic finding is a de novo nonsynonymous variant in *CUX1* (Cut-like homeobox 1) in an MZ twin pair (cases 1 and 2). The variant is novel, has a high CADD score (38), and is pathogenic according to AM and VarSome. Case 1 was typically developing until the sudden onset of neuropsychiatric symptoms that occurred following a documented group A beta hemolytic *Streptococcus* (GAS) infection. Symptoms met PANS criteria, including abrupt onset and severe eating restriction, oppositional behavior, aggression, reduced speech, cognitive decline, psychotic symptoms, incontinence, and self-harm. Case 2 is the MZ twin who, unlike case 1, was diagnosed with ASD as a child. The parents began to notice a significant worsening of behavior following infections, one of which coincided with the GAS infection detected in Case 1. Symptoms included separation anxiety, regression in speech, and refusal to put on clothes or leave the house. Another episode caused a regression in emotional regulation, impulse control, and cognitive ability.

Case 3 has a nonsynonymous, ultrarare variant in *CUX1* that is viewed as pathogenic by AM and VarSome. Case 3 has a history of Periodic Fever, Aphthous Stomatitis, Pharyngitis, and Adenitis (PFAPA), an autoinflammatory disorder, and exhibited infection-triggered psychiatric symptoms in childhood consistent with PANS (see Discussion).

*CUX1* codes for a transcription factor that has been implicated in NDD. It plays a role in DDR by base excision repair and interacting with ATM and regulates the expression of cytokines and chemokines [54–58].

### Case 4: *STK19* and *UVSSA*

Case 4 is a child with recurrent infections and PANS symptoms who was also diagnosed with enthesitis-related arthritis (ERA). He also has a history of recurrent aphthous ulcers and chronic gastrointestinal discomfort that has not been investigated which improved, along with PANS and ERA with Humira.

Splice disrupting variants were found in two DNA repair genes, STK19 and UVSSA, which act synergistically [59–61]. *STK19* was one of several genes found in a genome wide association study (GWAS) in metabolic syndrome and inflammation, and *UVSSA* is one of 27 genes identified in a natural killer cell transcriptome-wide association study carried out across five autoimmune disorders [62, 63].

Case 4 also has a pathogenic splice donor mutation in *DUOX2,* which codes for a member of the dual NADPH oxidase subfamily that generates hydrogen peroxide and is critical for the production of thyroid hormone [64, 65]. It also plays a role in host defense and chronic inflammation, in part through an interaction with IL-17 signaling, which contributes to gut barrier immunity [66, 67]. We have identified two other PANS and regression cases with pathogenic *DUOX2* variants (unpublished observations). The potential impact of *DUOX2* on PANS and regression through an effect on gut barrier immunity will be described in the Discussion section. All secondary variants are described in detail in **Supplemental Table 2**.

It should be noted that case 4 has a sibling with ERA and Undifferentiated Systemic Autoinflammatory Disease who neither carries the *DUOX2, STK19*, nor *UVSSA* variants, but instead has variants in *NOD2* (c. C2104T p.Arg702Trp, rs2066844) and PSTPIP1 (c.C492G:p.Asn164Lys) that are absent in Case 4. Both are viewed as benign variants according to AM and VarSome. However, *PSTPIP1* Asn164Lys has never been described, and the gene, which codes for a cytoskeleton-associated adaptor protein involved in pyrin inflammasome formation, has been implicated in autoinflammatory disorders [68, 69]. *NOD2* Arg702Trp is one of the most commonly analyzed variants in autoinflammatory disorders and common variable immune deficiency (CVID) [68–72].

The divergent genetic profiles in these siblings with autoimmune/autoinflammatory disorders are consistent with the bilineal family history of these disorders in case 4 in the context of shared environmental exposures.

### Case 5: *MYT1*

Case 5 is a child who met diagnostic criteria ASD and PANS. There is no history of developmental delays or issues with sociability or verbal communication. However, there is a history of hyperactivity, hypersensitivity to sound/light/touch, anxiety and emotional lability.

Following a viral infection, the child developed abrupt-onset OCD, restricted eating, separation anxiety, panic attacks, nightmares, enuresis, cognitive decline and regression in handwriting skills.

A pathogenic nonsynonymous variant was found in *MYT1,* which codes for a zinc-finger transcription factor involved in DNA repair during the G2/M checkpoint [73, 74].

Microduplications of the gene are inherited as a dominant trait with variable penetrance and result in a variety of neuropsychiatric problems including developmental and speech delays, ASD, mild-to-moderate intellectual disability, schizophrenia, and behavioral disorders [75].

### Case 6: ***TEP1***

Case 6 was diagnosed with ASD as a child and soon thereafter experienced his first episode of infection-induced OCD, restricted eating, enuresis, and mutism with echolalia. Psychiatric symptoms improved coincident with azithromycin, but mutism persisted with minimal improvement over time. Several years later, infection-induced behavioral decline occurred again, along with self-harm and refusal to walk. Bilateral calcification of the basal ganglia was found on MRI. There was a good response to an immune modulator, with improved social interactions, but mutism has persisted.

A predicted splice acceptor loss was found in *TEP1,* which codes for telomerase protein component 1, a component of the ribonucleoprotein complex responsible for telomerase activity, which is critical in maintaining genomic stability [76, 77]. TEP1 is also a component of the vault complex, a combination of proteins and non-coding RNAs (vault RNAs) that plays a role in synapse formation [76–78].

Several other variants were found that are of interest in this case because of their connection to DNA repair, neurodevelopment, immune function, and possible involvement of the gut. A stopgain mutation was found in *BTNL9*, a member of the butyrophilin family that plays a role in inflammatory bowel disease (IBD) [79, 80], and a novel frameshift mutation was found in *VNN1*, which affects the gut mucosal barrier [81–83].

Case 6 has a sibling with PANS who also developed infection-induced catatonia, who has the *TEP1, BTNL9* and *VNN1* variants.

### Case 7: USP45/PARP14

Case 7 is a child with autoimmune neutropenia who had a neuropsychiatric deterioration that met PANS criteria following an infection. The worst episodes occurred after SARS-CoV-2 infections. The child met developmental milestones, but had sleep dysfunction, emotional outbursts, and oppositional behavior. There is also a history of recurrent abdominal discomfort, diarrhea, and nausea but gastroenterology evaluation was negative.

We found a stopgain mutation in *USP45* that was scored as pathogenic by AM and likely pathogenic by Franklin (PVS1). USP45 is a deubiquitylase that acts as an oncogene by regulating DNA repair and immune checkpoint signaling through an association with ERCC1, a subunit of the DNA repair endonuclease XPF-ERCC1 [84–86]. XPF-ERCC1 DNA repair defects trigger neuroinflammation and neuronal cell death [87].

A disruptive splice donor mutation in *PARP14* was also found. PARP14 is involved in DNA repair via a CHK1-ATR pathway [88]. It also regulates innate immune function via IL-4 and IFN-γ signaling pathways [89] and altered expression has been found in five different genome wide expression datasets in Systemic Lupus Erythematosus (SLE) [90].

A compelling additional mutation we found was a de novo frameshift mutation in *CCR9*, which codes for the chemokine CCL25 receptor. CCL25 affects gut homeostasis permeability and will be described in greater detail in the Discussion [91–93].

### Case 8: *EP300*

Case 8 was diagnosed with ASD in early childhood following an evaluation for speech delay, poor eye contact, and repetitive behaviors. Several years later, there was abrupt onset of symptoms that met PANS/PANDAS criteria (i.e. followed a [GAS] infection). Since then, there have been multiple infection-induced flares. Following a Coxsackievirus infection, severe exacerbation of self-harm, aggressive behavior, and eating refusal resulted in hospitalization. The child is now non-verbal, although baseline language skills were minimal.

Case 8 has a haplotype consisting of two ultrarare variants in *EP300,* which codes for a histone acetyl transferase chromatin modifier that facilitates DNA repair and the transcriptional activation of DNA repair genes [94–96]. The primary variant is a novel nonsynonymous mutation with a CADD score of 24.9, which was classified as pathogenic by AM but was considered a VUS by VarSome and ClinVar. The other is an in-frame deletion that was also found in a patient with Rubinstein-Taybi syndrome (de novo in that case), an autosomal dominant NDD that also leads to immunodeficiency and autoimmune problems [97–99]. A different de novo nonsynonymous variant was also found in a patient with infection-induced neuropsychiatric decompensation [50].

An ultrarare, nonsynonymous variant was also found in the *CSMD1* gene that was pathogenic according to AM. *CSMD1* codes for a brain-expressed, complement inhibitor that has been implicated in ASD and schizophrenia most likely through the effects of the complement system on synaptic remodeling [100–102].

### Case 9: *PIDD1*

Case 9 developed a very abrupt onset of OCD, depression, anxiety, and insomnia that met PANS criteria, precipitated by a GAS infection that occurred at a time of extreme emotional stress. The initial episode occurred as a teenager. Subsequently, there were several other episodes of tonsilitis and GAS pharyngitis over a two-year span associated with exacerbation of OCD and other neuropsychiatric symptoms followed by the emergence of Periodic Limb Movement Disorder, severe persistent fatigue, and attention deficit hyperactivity disorder (ADHD). These symptoms have persisted in adulthood, although the OCD symptoms have improved with standard treatment.

A likely pathogenic frameshift mutation was found in *PIDD1* (p53-induced death domain protein 1), which is involved in the DNA damage response pathway through interactions with p53 and FANCI [103, 104]. The frameshift mutation would remove the C-terminal death domain, which is involved in the regulation of apoptosis and inflammation through the activation of caspases and NF-κB [91–93, 105]. *PIDD1* Variants have been found in ADHD and an autosomal recessive NDD with psychiatric features [106, 107].

### Case 10: *TREX1*

Case 10 is a child with ASD who developed a profound neuropsychiatric decline following an infection (catatonia with mutism, restricted eating, behavioral and developmental regression, irritability, emotional lability, reduced socialization, and self-harm). These episodes are recurrent, lasting several weeks to months with gradual recovery.

A pathogenic variant was found in the *TREX1* gene, which codes for a DNA repair enzyme through its 3’>5’ DNA exonuclease activity. Mutations in the gene have been found in many autoimmune/autoinflammatory disorders and type 1 interferonopathies, including Aicardi Goutières Syndrome (AGS), Familial Chilblain Lupus (FCL), Retinal Vasculopathy with Cerebral Leukodystrophy (RVCL), SLE, and neuropsychiatric SLE [108] [109–112]. These disorders can be caused by homozygosity, heterozygosity, or compound heterozygosity. There is a report of a patient with SLE caused by heterozygosity of the same variant we found [113]. LOF *TREX1* mutations activate the cGAS-STING type 1 interferon pathway [114, 115].

A likely pathogenic, ultrarare variant was also found in *SLC7A7,* an amino acid transporter that is highly expressed in peripheral blood leukocytes and small intestines that acts as an inflammation inhibitor [116, 117].

### Case 11: *ADNP*

Case 11 was diagnosed with high-functioning ASD and ADHD as a child and has a history of recurrent throat and sinus infections. She regressed during puberty with the loss of handwriting and reading skills. She also had severely restricted eating resulting in significant weight loss, contamination fears, anxiety, irritability, self-harm, behavioral regression, depression with suicidal ideation, chronic fatigue, and concentration problems. WGS revealed a nonsynonymous mutation in the *ADNP* gene that was scored pathogenic by AM and Varsome and has a CADD score of 28, but is viewed as a VUS by Franklin. However, the functional significance of this variant is supported by the presence of an ADNP-associated physical trait, hallux varus anomaly, that was also seen in the parent who transmitted the mutation. That parent has a history of inflammatory bowel disease and several cancers.

ADNP regulates DNA repair by resolving R-loops, which are three-stranded nucleic acid structures that accumulate on chromatin and contribute to genome instability, and by binding to POGZ, a regulator of homology directed repair [118, 119]. Genetic variants in both *ADNP* and *POGZ* cause ASD and the two have overlapping functions in a mouse model [120–122]. In addition, *ADNP* is one of the top ASD-associated risk genes associated with regression [123].

### Case 12: *SAMHD1*

Case 12 is a boy diagnosed with ASD and ADHD, who has infection-associated behavioral and psychiatric deteriorations including an episode of suspected autoimmune encephalitis. An ultrarare, pathogenic stopgain variant was found in *SAMHD1*, which codes for a dNTP triphosphohydrolase, an enzyme that plays a role in DNA repair, immune activation, and protection against HIV and other viruses by inhibiting their replication in immune cells [124, 125]. *SAMHD1* inhibits cGAS-STING and NF-κB, and LOF mutations cause Aicardi-Goutières Syndrome (AGS), an autosomal recessive monogenic type I interferonopathy [126, 127].

An important additional genetic finding in this case is a pathogenic splice donor mutation in *GRIN2A,* a well-established epilepsy and ASD candidate gene, especially regressive ASD [35, 128].

### Case 13: *FANCD2*

Case 13 was diagnosed with PANDAS following multiple infections with GAS. Prior to that, she was a typically developing child who had academic success, although there is a history of frequent infections with GAS, upper respiratory infections, and urinary tract infections. Acute behavioral decline characterized by auditory hallucinations, debilitating intrusive thoughts, restrictive eating, and coughing tic occurred following infections other than GAS. She was subsequently diagnosed with schizoaffective disorder and hypothyroidism.

An ultrarare pathogenic stopgain mutation was found in *FANCD2* a member of the FAC family that plays a crucial role in DNA repair and R-loop regulation, particularly in responding to DNA damage caused by interstrand crosslinks [129, 130]. In addition, it also functions as a regulator of ferroptosis, which is a key factor in the development of autoimmune diseases including SLE, Rheumatoid Arthritis (RA), IBD, and Multiple Sclerosis (MS) [131, 132].

Case 13 has a sibling who has been diagnosed with PANS but carries a pathogenic variant in a different DNA repair gene, *RNASEH2B* (Ala177Thr; pathogenic/likely pathogenic according to ClinVar and pathogenic by Franklin, PM3 criteria). This variant is commonly found in AGS [133] (see Discussion). WES is pending in the sibling. Like case 4, the divergent genetic profiles could be due to the strong bilineal family history of autoimmune and autoinflammatory disorder found in this family along with shared environmental exposures.

### Case 14: *RAD54L*

Case 14 has a history of abrupt onset of OCD, tics, anxiety, oppositional behavior, and cognitive decline lasting days to several weeks. These generally occur following infections, including GAS, but most have not been well-characterized. Prior to the first flare, there is a history of delayed speech and a learning disability, especially in math. There was no response to antibiotics nor IVIg. In fact, a transient worsening of symptoms was reported by the parents after IVIg. Multiple flares recurred over a period of 4 years, but episodes have been milder since the onset of puberty. Similar to the prior reported cases, there is a strong history of autoimmune and autoinflammatory disorders on both sides of the family.

Case 14 inherited a stopgain mutation in *RAD54L* that was scored as likely pathogenic by Franklin. *RAD54L* codes for a member of the DEAD-like helicase superfamily that plays a role in homologous recombination and repair of double-stranded DNA breaks [134]. Although there are no reports of any connections between RAD54L and either neurodevelopmental or immune/autoimmune disorders, the gene is expressed in the brain and immune cells, with the highest levels found in EB-transformed lymphoblasts (GTEx database, Release 6).

Case 14 also carries an ultrarare frameshift deletion in *BRMS1*, which codes for a transcriptional repressor that has effects on DNA repair by regulating cell sensitivity to DNA interstrand crosslink damage through an interaction with FANCI [135]. It also functions as a regulator of NF-κB expression [136, 137].

### Case 15: *PRKN*

This is the first of two cases (cases 15 and 16) where the primary genes of interest affect mitochondrial function, in particular mtDNA replication and mitophagy, disruption of which can lead to the release of mtDNA into the cytoplasm, a potent inducer of cGAS-STING signaling [138–140]. Case 15 is a child with developmental delay and behavioral/psychiatric decline that met PANS criteria, who has a de novo PRKN exon 2/3 deletion. The child also suffered from severe gastrointestinal reflux resulting in poor weight gain in the first year. Reflux has required ongoing chronic acid suppression. Imaging studies revealed a thin corpus callosum and periventricular leukomalacia, a condition associated with NF-κB activation and experimentally induced neuroinflammation [141, 142].

*PRKN* codes for Parkin, which along with PINK1 regulates mitophagy [143–145]. Biallelic LOF mutations in *PRKN* causes early onset Parkinson’s disease (PD) [146]. Heterozygosity for partial *PRKN* deletions has been found in children with ASD [146–148].

An additional variant of interest we found was a stopgain mutation in *LGALS4* (Galectin-4). This gene is expressed almost exclusively in epithelial cells in the transverse colon and terminal ileum. LGALS4 and other galectins are involved in the development of IBD by regulating T-cell function in the gut and affecting host-gut microbe interactions [149, 150].

### Case 16: *POLG*

Case 16 is a child with ASD and catatonia who developed acute eating refusal and was diagnosed with Avoidant/Restrictive Food Intake Disorder (ARFID). Over several months, this progressed to a catatonic state characterized by persistent eating refusal and mutism.

Treatment with benzodiazepines and an antipsychotic increased alertness but provoked agitation and combativeness, without improving eating behavior or speech. Workups for autoimmune encephalitis, infections, and metabolic disorders were negative. There was no immediate response to antibiotics, antivirals, steroids, IVIg, and plasma exchange. However, over the course of approximately six months, she improved and was able to return home.

A novel, de novo nonsynonymous variant was found in *POLG,* which codes for a DNA polymerase responsible for replication and repair of mtDNA [151, 152]. Mutations in *POLG* result in host of autosomal dominant or recessive mitochondrial disorders that can cause psychiatric symptoms and cognitive decline [151, 153, 154]. The *POLG* variant is a VUS according to the Franklin database although it was given modest score for pathogenicity based on the variant’s very low MAF. The CADD score is 23. However, we included the variant in this study because it is novel and de novo, and its function as an mtDNA polymerase can be linked directly to innate immune activation; impaired mtDNA repair can lead to the release of mitochondrial DNA into the cytosol and activation of cGAS-STING and NLRP3 inflammasomes [138–140]. It is also interesting to note that the mutation overlaps with the 3’-UTR of the DNA repair gene, *FANCI* (*POLG* and *FANCI* are transcribed in opposite orientations and overlap in their 3’ ends). We previously reported a patient with PANS and seronegative autoimmune encephalitis who has a nonsense mutation in *FANCI*, a component of the FAC that repairs DNA interstrand crosslinks [1, 155]. How this variant affects the regulation of FANCI is not known.

Case 16 also has a de novo pathogenic variant in *PRRT2*, a regulator of neurotransmitter release associated with a variety of neurological and neurodevelopmental disorders. We hypothesize that the modest predicted effect of the *POLG* variant is acting synergistically with dysfunctional neurotransmitter release to cause this child’s severe neuropsychiatric episode.

## Protein–Protein Interaction Network Analysis

To further explore the biological relationships among the DNA repair gene variants, we performed a STRING-based protein-protein interaction (PPI) analysis using all genes from this and our previous two studies [1, 38] (**Figure**). The interacting genes are arranged according to biological function. The confidence level of the interactions is represented by the thickness of lines between the proteins (see methods).

## Discussion

This study expands on our previous findings that link mutations in genes involved in DDR to complex cases with PANS, regression in ASD or other NDDs [1, 38]. The findings are also supported by recent genetic studies in acute-onset neuropsychiatric disorders, and DSRDS [49, 50]. Although the functions of the genes identified in these two studies highlighted those that affect type I interferon signaling and chromatin regulation, the majority also affect DNA repair.

The overlap between our respective studies is highlighted by the finding of common genes, e.g., *EP300* in the Dale et al. study and case 8. In addition, Jafarpour described a de novo mutation in *RNASEH2A*, which is one of the genes, along with *SAMHD1* (case 12) and *RNASEH2B* (sibling of case 13) that can cause Aicardi-Goutières Syndrome (AGS), a type 1 interferonopathy, autoinflammatory disorder [126]. Although AGS is typically inherited as an autosomal recessive disorder, these findings suggest that heterozygotes have an increased risk of developing infection-induced neuropsychiatric decompensation, most likely in the context of additional genetic variants.

An association with an autoinflammatory disorder was also seen in case 3 who was diagnosed with PFAPA. Although variants in *CUX1* have not been described in PFAPA, DNA damage and oxidative stress in peripheral blood mononuclear cells do occur [156]. In addition, we have a sibling pair and a singleton with PANS who have been diagnosed with Familial Mediterranean Fever (FMF), each of whom carries variants in the *MEFV* gene associated with this condition (R202Q and A744S, unpublished observations). These findings suggest that some PANS cases have an underlying autoinflammatory disorder and that physicians caring for FMF and PFAPA children should be aware that neuropsychiatric issues could have an immune/inflammatory-based etiology requiring unique diagnostic and treatment strategies.

In our previous papers [1, 38] and the current paper, we collected clinical data and analyzed WES from unselected families, only a few of whom had sequencing carried out on trios while the Jafarpour et al. and Dale et al. papers used family trios to identify de novo variants. The different strategies have advantages and disadvantages. We have confirmed de novo variants in DNA repair genes in *PPM1D* (3 cases) [1, 38] and in *CUX1, POLG* and *PRKN* in the current study. On the other hand, trio analysis to detect de novo variants may have limitations in our cohort given the extraordinarily high rate of autoimmune and autoinflammatory disorders in patients and their families, which suggests transmitted rather than de novo variants. This high rate of comorbidity between neuropsychiatric decompensation and family history of immune disorders may be due to selection bias. This is especially true for the variants found in case 13 who has an affected sibling with a different pathogenic candidate variant in *RNASEH2B*, which affects DNA repair and innate immunity.

Researchers have suggested that autoimmune and autoinflammatory processes underlie the development of behavioral regression in PANS and in a subgroup of individuals with ASD [36, 157–160]. Our findings highlight impaired DDR as a potential mechanism for these associations. Impaired DDR can lead to leakage of nuclear DNA into the cytosol, which can trigger the cGAS-STING/ type I interferon antiviral innate immune response — especially following an infectious disease event, which activates the same pathway and heightens cellular stress and DNA damage [47, 161–163]. Non-infectious cellular stressors such as cellular injury, heat shock, and oxidative stress can also induce DNA damage and activate innate immune signaling [164, 165]. AIM2 inflammasome signaling, which leads to the induction of IL-1β and IL-18, is also activated by cytosolic DNA [166, 167]. DNA repair deficits can also lead to an increase in reactive oxygen species, which can activate NLRP3 inflammasomes [168, 169].

In addition to these canonical connections between DDR and innate immunity, faulty DNA repair can also disrupt autophagy, apoptosis, and mitochondrial function, and cause lysosomal damage and cell senescence [170, 171].

Although physiological activation of cGAS-STING is central for the response to foreign viral DNA and RNA [172], studies show that over-activation can lead to autoimmune, autoinflammatory, and neurodegenerative disorders [173–176]. For example, increased IFN-I responses mediated by cGAS-STING due to abnormal clearance of cytosolic DNA and RNA have been implicated in the pathogenesis of SLE and aggressive rheumatoid arthritis [177–180]. Circulating type I IFN levels are elevated in approximately 50% of patients with SLE and IFN receptor inhibition is an approved therapy [181, 182]. A substantial fraction of SLE patients have neuropsychiatric symptoms [183, 184]. It has also been proposed that cGAS-STING signaling plays a role in neurodegenerative disorders, including Alzheimer’s Disease, PD, Amyotrophic Lateral Sclerosis, Huntington’s disease, and MS [185–187]. Markers for neurodegenerative disorders are elevated in 27% of patients with psychiatric disease preselected for suspected immunological involvement and phenotypes including catatonia, agitation, and acute onset of symptoms similar to the cases described here [188]. Early immunological interventions may mitigate some damage, but this needs to be confirmed in clinical trials. However, other forms of damage may accumulate over time that are not responsive to immunological interventions.

In addition to nuclear DNA, disruption of mitochondrial integrity can also lead to the release of mitochondrial DNA into the cytoplasm and innate immune activation, as noted above [138–140]. *The PRKN and POLG* de novo variants described in this report affect mitochondria through disrupted mitophagy and mtDNA synthesis, respectively. It is interesting to note that some FAC proteins also regulate mtDNA replication fork stability, which can activate the mtDNA-dependent cGAS/STING response when dysfunctional. For example, FANCI has been found to regulate PRKN-mediated mitophagy [189, 190]. We previously identified a nonsense mutation in *FANCI* in a patient with PANS and seronegative autoimmune encephalitis [1]. In addition, FANCD2 (case 13) has non-canonical effects on mitochondrial function [191].

It is also interesting to place our DNA repair-related findings in the context of molecular and genetic studies carried out in OCD. For example, levels of a biomarker for oxidative stress and DNA damage, 8-hydroxy-2’-deoxyguanosine (8-OhdG), were significantly higher in OCD patients than in controls, while 8-OHdG levels were significantly lower in treated patients than in those who have been newly diagnosed [192]. And several genetic studies show that a subgroup of OCD patients has mutations in genes that affect the DDR, including the well-established DNA repair regulator, BRCA2 (FANCD1) [193, 194].

Finally, an interesting additional finding in our study is the detection of likely pathogenic variants in several genes that can affect the gut microbiome/homeostasis, including *BTNL9, VNN1, LGALS4, DUOX2,* and most notably a de novo variant in *CCR9* (case 7). CCR9 is the receptor for the chemokine CCL25. CCR9/CCL25 forms a signaling axis critical for T-cell homing to the small intestine that has been implicated in gut immune surveillance, IBD, gut innate immunity, and autoimmune disorders [195–197]. *DUOX2* codes for dual oxidase 2, an enzyme that produces hydrogen peroxide in the gut. DUOX2 is upregulated in response to microbial dysbiosis or TLR4 signaling, and genetic and molecular studies show that it is involved in IBD [198–200]. In addition to case 4, we have identified two other PANS cases with pathogenic mutations in *DUOX2* (unpublished observations). We propose that genetic variants affecting gut microbiome balance may contribute to neuroinflammation and behavioral symptoms by altering neuroimmune and metabolic pathways. Microbial metabolites or immune-modulatory molecules produced in the gut could cross biological barriers and influence brain function or provoke neuroinflammatory responses [201–203]. A comprehensive assessment of all variants that are expressed primarily in the gut found in our cohort that might impact the gut microbiome is beyond the scope of this paper.

Clinically, if our findings regarding genetic vulnerability in DNA repair are replicated in larger and independent cohorts, they may eventually inform the development of more specific diagnostic tools and targeted interventions for infection-associated psychiatric episodes and behavioral decompensation, potentially by modulating overactive immune pathways such as type I interferons and AIM2 inflammasomes. However, our results should be interpreted with care. Independent validation is essential, including integration of longitudinal clinical data, comprehensive genetics, and immunophenotyping. This caution is warranted for all genetic studies given the inherent complexity and multifactorial basis of inheritance and is especially pertinent due to our study’s small sample size and the use of genetic data from multiple sources, which may introduce technical and analytical biases. To address these limitations, a whole-exome sequencing (WES) of a larger cohort with PANS cases is currently underway. In parallel, functional analyses using patient-derived immune cells and microglia from induced pluripotent stem cells (iPSCs) are ongoing. These efforts aim to clarify the mechanistic underpinnings of our observations and to refine future hypotheses.

## Supporting information

supplemental table 1 expansion of table

supplemental table 2 additional genes

## Data Availability

National Database for Genomic Data (NDA): A repository for human subjects data collected from research projects

## Acknowledgments

The authors want to thank the participating families.

## Statement of Ethics

Written informed consent was obtained from each parent of participants prior to the study. The consent form was approved by the Albert Einstein College of Medicine (protocol 2022-14636) and was administered by the corresponding author.

## Author contributions

D.V and A.S. validated variants, generated the String analysis, and contributed to the manuscript; R.A.D processed all WES data; E.P. processed DNA, validated variants, carried out AlphaMissense, and copy edited the manuscript; A.S. contributed a case; J.S.L. contributed a case; I.P.C. contributed to the evaluation of the POLG variant; J.M. contributed a case; S.M.A.S. analysis of the ADNP variant; P.J.S. identified the ADNP variant; J.F. contributed cases and contributed to the manuscript; J.L.C. contributed to manuscript; H.M.L. study design, wrote manuscript, interviewed families. All authors reviewed the manuscript.

## Data availability statement

N/A

## Supplemental files

**Supplemental methods.** Additional methods and primers used for validation

**Supplemental Table 1.** Comprehensive assessment of each gene shown in the main Table. Abbreviations: Age at onset of symptoms given in blocks of 5 years; (MAF (minor allele frequency); chr. (Chromosomal locus, hg19 coordinates); rs # (Reference SNP cluster ID); CADD score (Combined Annotation Dependent Depletion – see methods for details); AM (AlphaMissense).

**Supplemental Table 2.** List of additional variants of interest. See Table Legend for abbreviations

---

**Figure:**
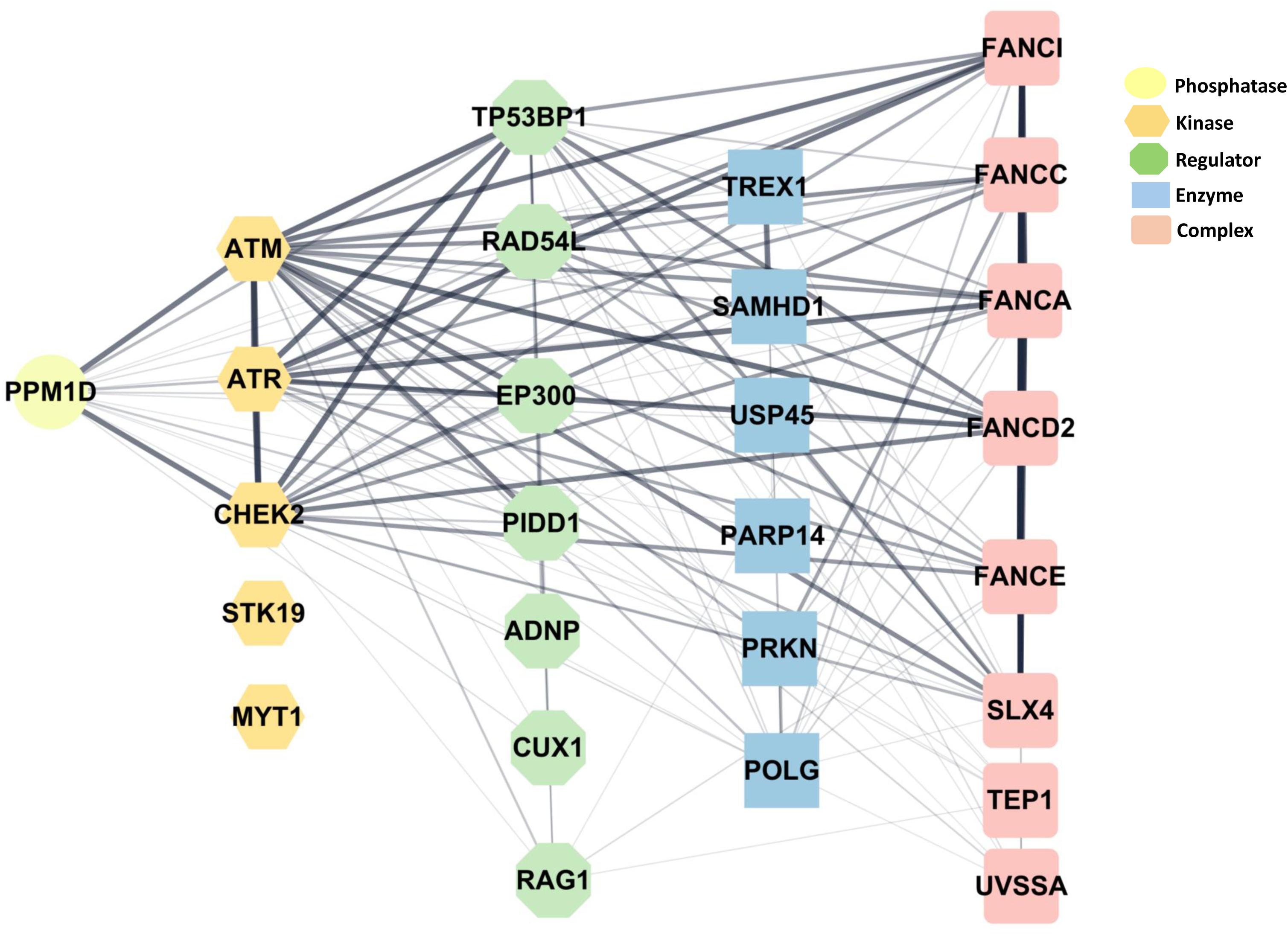
Protein–protein interaction (PPI) analysis. Shows relationships among the DNA repair and mitochondrial genes described in this paper, along with those found in our previous studies [1, 38]. See methods section for details

**Table.** DNA repair and mitochondrial gene variants. For a more comprehensive assessment of each variant, see Supplemental Table 1.

**Table: DNA Repair and Mitochondrial Variants**
| Case# | StudyID | Diagnosis | Sex/Age at Onset | DNA repair genes | Variant |
| --- | --- | --- | --- | --- | --- |
| 1 | DC_PANS | PANS/Cognitive Decline | F/6-10 | CUX1 | nonsynonymous, c.G1808A; p.R603Q |
| 2 | JN_AS DR | ASD/regression | F/1-5 | CUX1 | nonsynonymous, c.G1808A; p.R603Q |
| 3 | B1PFA | PANS/PFAPA | M/6-10 | CUX1 | nonsynonymous, c.T1822C;p.Y608H |
| 4 | DUZG | PANS | M-5-10 | STK19/UVSSA | splice donor, c.815+1G>A/splice acceptor, c.551-1G>T |
| 5 | MC_PANS | regression/PANS | F/1-5 | MYT1 | nonsynonymous, c.C2726T;p.A909V, C2807T |
| 6 | LM_AS DRP | ASD/regression | M/1-5 | TEP1 | splice acceptor exon5:c.871-2A>G |
| 7 | MKJ_PANS | PANS | F/6-10 | USP45/PARP14 | stopgain, A1462T; p.K488*/splice donor exon2:c.321+1G>A |
| 8 | OBF_PANS | ASD/regression/PANS | M6-10 | EP300 | nonsynonymous, c.C7043T; p.S2348F/in-frame del c2209-2213 |
| 9 | EPI24-NTEP | PANS | M16-20 | PIDD1 | frameshift c.2308delG; p.A770Lfs*8 |
| 10 | AF_PANS | ASD/regression | F/1-5 | TREX1 | c.G311A; p.R104H:c.G341A; p.R114H |
| 11 | SGAD | ASD/PANS/regression | F/11-15 | ADNP | nonsynonymous, c.2149C>; p.R717C |
| 12 | NF_PANS | ASD/infection-induced behavioral changes | M/1-5 | SAMHD1 | stopgain, c.658C>T; p.Arg220* |
| 13 | OA_PANS | PANS | F/6-10 | FANCD2 | stopgain,c.C2380T; p.R794* |
| 14 | GN_PANS | PANS/learning disability/dyscalculia | M6-10 | RAD%\$L | stopgain, c.C1525T; p.R509X |
| Case# | StudyID | Diagnosis | Sex/Age at Onset | Mitochondrial Genes | Variant |
| 15 | KDWP | ASD/PANS | F/1-5 | PRKN | exon 2/3 del |
| 16 | AMPP | ASD/seronegative AE | F/1-5 | POLG (FANCI 3'UTR) | nonsynonymous, c.3677T>A; p.L1226H |

