## supplemental methods for "Ultrarare Variants in DNA Damage Repair and Mitochondrial Genes in Pediatric Acute-Onset Neuropsychiatric Syndrome and Acute Behavioral Regression in Neurodevelopmental Disorders"

Saliva DNA was obtained using a DNA Genotek kit (cat# OGR-500). DNA was extracted using prepIT purifier (DNAgenotek cat# PT-L2P-5) according to the manufacturer’s instructions, except for the inclusion of a phenol/chloroform purification step (Phenol:Chloroform:Isoamyl Alcohol 25:24:1 saturated with 10mMTris pH8.0, 1mM EDTA; followed by the addition of 1ul glycogen, sodium acetate, pH 5.2 to a final concentration of 300 mM, and an equal volume of Isoamyl Alcohol to precipitate DNA). Genomic DNA was amplified using HotStarTaq DNA Polymerase (Qiagen cat# 203203) according to the manufacturer’s instructions using primers that flank the variants of interest (see list below). PCR products were purified using a QIAquick PCR Purification Kit (Qiagen, catalog# 28104) according to the manufacturer’s instructions. DNA was sequenced by the standard Sanger dideoxy chain termination method using one of the two PCR primers. The primers used are shown below.

**Primers used to validate DNA repair and secondary genes**

CUX1-F: ACCCTAGGGCCCTTTCTGT

CUX1-R: AGAGATGCAGCTTGGGGTAG

STK19-F: TGCGCTCAGATCAAGAATCCA

STK19-R: GCGGATTGGCTCCCTCCA

MYT1-F: GAGTTGGGCTCCCCTTGT

MYT1-R: GGTCGGTCCTTCATTCTTCA

RAD54L-F: AGGGCTGTCCTGTTTCAAGG

RAD54L-R: CCCCTTTGGCATCCTGGTTA

TEP1-F: CACCCTGTCTCTCTCCAAGC

TEP1-R: GACCAGAGTCAAGGGAGGTG

USP45-F: TAAGACCTCAAATGCTTCCC

USP45-R: GGTGATAAGGAAATGGCAGA

EP300-F: CACTCCAGTCCTTCCCCAAG

EP300-R: TGCTCCCAAAATACTACAAGGTGT

EP300new-F: CCAGGCTCAGCAGATGAACA

EP300new-R: CTGAAGGAGTCGCTGCTGAT

TREX1-F: AGACAAGCTCTCCCTGTGTG

TREX1-R: CCTTCAGCGCAGTGATGCTA

ADNP-F: GCGATGCCTTTTCATGCCAA

ADNP-R: CCCAATCATGGCAGTGACCT

FANCD2-F: CCTCCAGGTTTTATTGGCTTGC

FANCD2-R: GGGGGTTAGTGCTTGGTGAA

RIPK2-F: TCCTAATCATCTCCAGTTAAAGTGT

RIPK2-R: TGGAGATTGACATGAGCTACCA

RNASEH2B-F: GGTCTGAAGGCCACCTATGA

RNASEH2B-R: GCATTTACACATAAGCAACTTACTCC

BTNL9-F: CCAAGCCCACACACGTCTTA

BTNL9-R: GCTCGGACAACCACAGATGT

VNN1-F: AATAGCGCCCTTCCACAGTG

VNN1-R: TCTCGCAACTGGATTCCCAC

CCR9-F: CCCTTGCAGAGCCCTATTCC

CCR9-R: AGAGGAGGTCAGCAATTGCC

CSMD1-F: AAATCCTTGCCGGTTTTGCG

CSMD1-R: GTCTGGGGAGACTGCTAACAC

SLC7A7 F: GGACTTAAGGATGCACGGCT

SLC7A7 R: AAGAAGGCTCAGTGCGCTAA

GRIN2A-F: GGACAGCAAGAGGAGCAAGT

GRIN2A-R: GTTTGTAAGGGTCCGAGGGG

NOD2-F: TCTTTGAGCACTGCTGTTGG

NOD2-R: GAACTCGGTGCGGATGTACT

RAD54L-F: AGGGCTGTCCTGTTTCAAGG

RAD54L-R: CCCCTTTGGCATCCTGGTTA

BRMS1-F: GATGTGGCTTTGACTTCGGC

BRMS1-R: GACCCTCAGAGCAGCTGG
